# Evolutionary trends of the COVID-19 epidemic and effectiveness of government interventions in Nigeria: A data-driven analysis

**DOI:** 10.1101/2020.05.29.20110098

**Authors:** Oluwakemi O Odukoya, Ismaila A Adeleke, Chris S Jim, Brenda C Isikekpei, Chiamaka M Obiodunukwe, Folusho E Lesi, Akin O Osibogun, Folasade T Ogunsola

## Abstract

**Background:** Nigeria became the first sub Saharan African country to record a case of COVID-19 after an imported case from Italy was confirmed on February 27, 2020. Moving averages and the Epidemic Evaluation Indices (EEI) are two important but complementary methods useful in monitoring epidemic trends, they can also serve as a useful guide for policy makers and inform the timing of decisions on preventive measures. The objectives of this paper are to graphically depict the trends of new COVID-19 cases nationally and in two key States (Lagos and Kano) and the Federal Capital Territory (FCT) using the moving averages and the EEI. In addition, we examined the effects of government’s public health interventions on the spread of COVID-19 and appraise the progress made so far in addressing the challenges of COVID-19 in Nigeria.

**Methods:** We used data on new cases of COVID-19 from public sources released by the Nigeria Center for Disease Control (NCDC) from the 27^th^ February 2020, when the first case was recorded, to 11^th^ May, 2020, one week after the lockdowns in Lagos and the FCT were lifted. We computed moving averages of various orders, the log transformations of the moving averages and then the EEIs of new COVID-19 cases for Nigeria as a whole, and then for two of the most affected states i.e. Lagos and Kano, as well as the Federal Capital Territory (FCT). Then, we plotted graphs to depict these indices and show the epidemic trends for COVID-19 in each scenario.

**Results:** Nationally, the number of new cases of COVID-19 showed an initial gradual rise from the first reported case on the 27^th^ February 2020. However, by the second week in April, these numbers began to show a relatively sharper increase and this trend has continued till date. Similar trends were observed in Lagos state and the FCT. The rate of growth of the logarithm-transformed moving average in the period leading to, and including the lockdowns reduced by a factor of 0.65. This suggests that the policies put in place by the government including the lockdown measures in Lagos and the FCT may have had a positive effect on the development of new cases of COVID-19 in Nigeria. Nationally and in Lagos, the EEIs of the COVID-19 cases started off on very high notes, however, the effects of the lockdown gradually became evident by the end of April and early May 2020, as the EEIs headed closer to 1.0. In all case scenarios, the EEIs are still above 1.0.

**Conclusions and recommendations:** The number of new cases of COVID-19 has been on a steady rise since the first reported case. In Nigeria especially across the two states and the FCT, public health interventions including the lockdown measures appear to have played a role in reducing the rate of increase of new infections. The EEIs are still above 1.0, suggesting that despite the progress that appears to have been made, the epidemic is still evolving and Nigeria has not yet reached her peak for COVID-19 cases. We recommend that aggressive public health interventions and restrictions against mass gatherings should be sustained.

## Introduction

The novel coronavirus (COVID-19) caused by severe acute respiratory syndrome coronavirus 2(SARS-CoV-2), first reported in China in December 2019, has been declared a global pandemic. There are over four million cases and more than 300,000 deaths worldwide.^1^

On the 14^th^ February, Africa’s first COVID-19 case was recorded in Egypt. Three months later, (May 13 2020), the disease had spread throughout every African country, with Lesotho being the last to record a new case.^2^ In Africa, there are a total of 86,721 cumulative confirmed cases, 2,787 deaths reported and 33,840 recoveries as at May 19 2020.^3^ Four countries alone, South Africa, Egypt, Algeria and Nigeria recorded 47% of the cases in Africa. Though Africa so far has the lowest number of cumulative confirmed cases, there are concerns regarding the representation of the true prevalence of the disease, as several countries report unavailable or low testing capacities. Furthermore, under-developed health systems and the existence of several hard to reach and/or highly rural areas, also pose a threat to Africa’s ability to contain and mitigate the spread of the disease and manage confirmed cases effectively.

Nigeria became the first sub Saharan African country to record a case of COVID-19 after an imported case from Italy was confirmed on February 27, 2020. As at May 18^th^ 2020, of the 36,899 tests carried out so far, there have been a total of 6,175 confirmed cases and 191 deaths in Nigeria, 1,644 cases have been discharged with 4,340 cases remaining active. Of the 36 states and the Federal Capital Territory Abuja, only two states in Nigeria (Cross Rivers and Kogi states) are yet to record any confirmed cases of COVID-19.^4^ It is important to note that these figures do not account for asymptomatic or untested cases.

The Nigerian government, as with other national governments are grappling with the rising numbers of COVID-19 cases and have instituted several containment measures in a bid to stem this rising tide. Various steps have been taken by both Federal and State Governments to curb the spread of the virus, including restriction of interstate movements, closures of schools, markets and other business places, bans on religious and social gatherings as well federally-ordered lockdowns in two states and the FCT. Other measures include social distancing, use of nose masks, good hygiene, proper hand washing and use of alcohol-based hand sanitizers, sneezing into elbows, etc. as advised by the Nigeria Center for Disease Control. However, considering the socio-economic well-being of citizens and the economic challenges that arise as a result of some of the public health measures, some of these restrictions are gradually eased in some parts of the country. This is amidst the rising number of confirmed cases and deaths, as well as the spread of the virus to previously unaffected states. Accordingly, there is need to evaluate the trends of the COVID-19 epidemic in Nigeria, vis-à-vis government’s public health response to contain and mitigate against the spread of the disease.

The study shows the evolutionary trends of the COVID-19 outbreak in Nigeria. We use two methods to monitor and predict the COVID-19 case trends nationally and in two of the most severely affected states as well as the FCT. Firstly, the moving average method, this helps to smoothen out the noise of random outliers and emphasize long-term trends. They are best visualized using simple plots and are useful for showing epidemic trends over time.^5^ Secondly, we compute the Epidemic Evaluation Index (EEI) which has been used successfully in China to monitor COVID-19 trends.^5^ The EEI serves as a useful tool to monitor and predict the evolutionary trends for the emergence of new COVID-19 cases. This index was developed by He He, Wang and Wang (2020) as an alternative to traditional epidemic prediction models that are highly sensitive to the pre-specified epidemiological parameters and tend to have low reliability in the face of the possible seasonality of coronavirus, inter-city mobility, transmission of travel behavior related transmission and the number of asymptomatic infections.^5,6^ The traditional model also has the problem of underestimation of the scale of outbreak.^6^ The EEI aims to comprehensively analyze the epidemic situation based on combination of actual observation and log of moving averages (LMA) and can more accurately reflect the true situation of the epidemic. This index could be used to support the decision-making process in epidemic control and prevention strategies. An Epidemic Evaluation Index of less than 1 is often preferable as this indicates that the epidemic is under control. Higher indices show higher rates of on-going disease transmission.^7^ The Epidemic Evaluation Index gives an indication of the rate of the escalation of new cases in an epidemic situation and serves as a useful tool to support the decision-making process for epidemic control and prevention strategies.

The objectives of this paper are two-fold. Firstly, we aimed to use the moving averages and EEIs to graphically depict the trends of new COVID-19 cases nationally and in Lagos, Kano and the FCT. Secondly, we examined the effects of some of the government’s public health interventions on the spread of COVID-19 and appraise the progress made so far in addressing the challenges of COVID-19 in Nigeria.

## Methods

### Data source

The data used in this study are obtained from public sources made available by the Nigeria Center for Disease Control (NCDC). From the first reported case of COVID-19 in Nigeria, the NCDC releases data on the number of new cases, number of discharges, number of deaths and the number of confirmed cases by state on a daily basis. More recently, the number of tests carried out also released. These data are available in the daily situation reports available at www.ncdc.gov.ng. The data used for this study date from February 27, when Nigeria recorded her first COVID-19 case to May 11, one week after the lockdown in Lagos and the FCT was eased.

Using this data, we computed moving averages of various orders, the log transformations of the moving averages and then the EEIs of COVID-19 cases, for Nigeria as a whole and then for other states i.e. Lagos, the epicenter of the epidemic in Nigeria, Kano and the FCT. Then, we plotted graphs to depict these indices and show the epidemic trends for COVID-19 in each scenario.

## Moving averages

Using the Moving Average and Moving Averages of Logarithm – Transformed Data, we computed moving averages of order 5, order 7, order 10 and order 14. The choice of the order of the moving average was based on the transmission dynamics as new cases are often expected to show symptoms up to 14 days after an infection. We used the 5-day, 7-day, 10-day and 14-day moving averages of the actual and log-transformed number of daily covid-19 cases. A moving average of a certain order simply means the average of the number of new cases of that order. For instance, a moving average of the order of 5, computes the average number of new COVID-19 cases over the past 5 days. We added 1 to the actual number of observations before logarithm transformation to eliminate the problem of having a zero entry on some days, especially when the test results are delayed or lagged. Transformation makes the index more tolerant of outliers and also brings better stability.^7^ If the number of new confirmed cases at time *t* is denoted by *X_t_*, then the moving average of order *n*, MA(n) is computed as

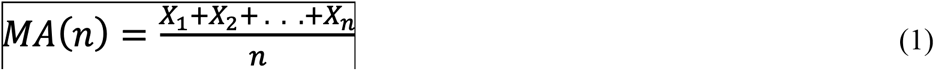

In epidemic trend analysis, a moving average helps smooth out the noise of random outliers and emphasizes long-term trends. Moving averages are usually plotted and best visualized. The averages are computed and visualized in the figures below. The logarithmic transformation of the observed new COVID-19 cases at time *t* is achieved by using:

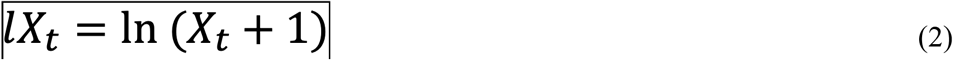

The moving average of the logarithmic transformed observed cases at time *t* is

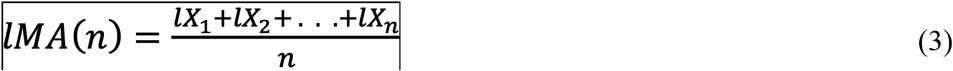

### Epidemic Evaluation Index (EEI)

The Epidemic Evaluation Index, as introduced by Hu, Hu, Wang and Wang, (2020) is defined as:

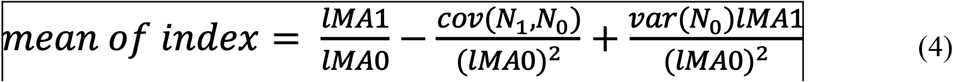

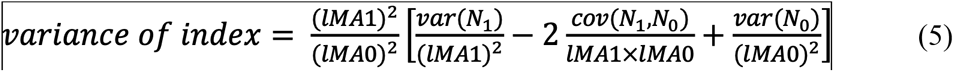

Where

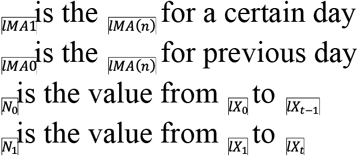

The computed EEI indicates the current epidemic situation in each scenario. The upper and lower limits represent the worst and the best estimates of the epidemic situation, respectively.

## Results

Nationally, the number of new cases of COVID-19 showed an initial gradual rise since the first reported case on the 27^th^ February 2020. However, by the second week in April, these numbers started to show a relatively sharper rise and this trend has continued till date. The highest number of new cases (381 new cases) were reported on the 8^th^ May, four days after the lockdowns in Lagos and the FCT were eased. Similar trends were observed in Lagos state. New cases in the FCT also followed a similar trend, but started to rise a little later i.e. by mid-April. In Kano, the first COVID-19 case was reported on the 11^th^April and started to show a sharp rise on the 24^th^ April (about two weeks later). Our analysis suggests that new COVID-cases in Nigeria may not yet have reached its peak, as the reports of new cases are still on the increase. See Figures 1-4 The rate of increase of new cases nationally began to rise sharply from the 18^th^ March, however, by the middle of April 2020, this sharp rise appeared to steepen a little. The rate of growth of the logarithm-transformed moving average in the period leading to, and including the lockdowns was estimated to be 0.08. However, the observed rate for the period after the lockdown is 0.052. Considering the incubation period of up to 11.5 days: 95% CI 8.2-15.6,^8^ there is often a time lag between the institution of government policies and the time when policy effects start to kick in. Our analysis suggests that the policies put in place by the government including the lockdowns in Lagos and FCT may have had a positive effect on the development of new cases of COVID-19 in Nigeria. The rate of rise of new cases reduced by a factor of 0.65. See Figures 5-8.

**Figures 1-4:**
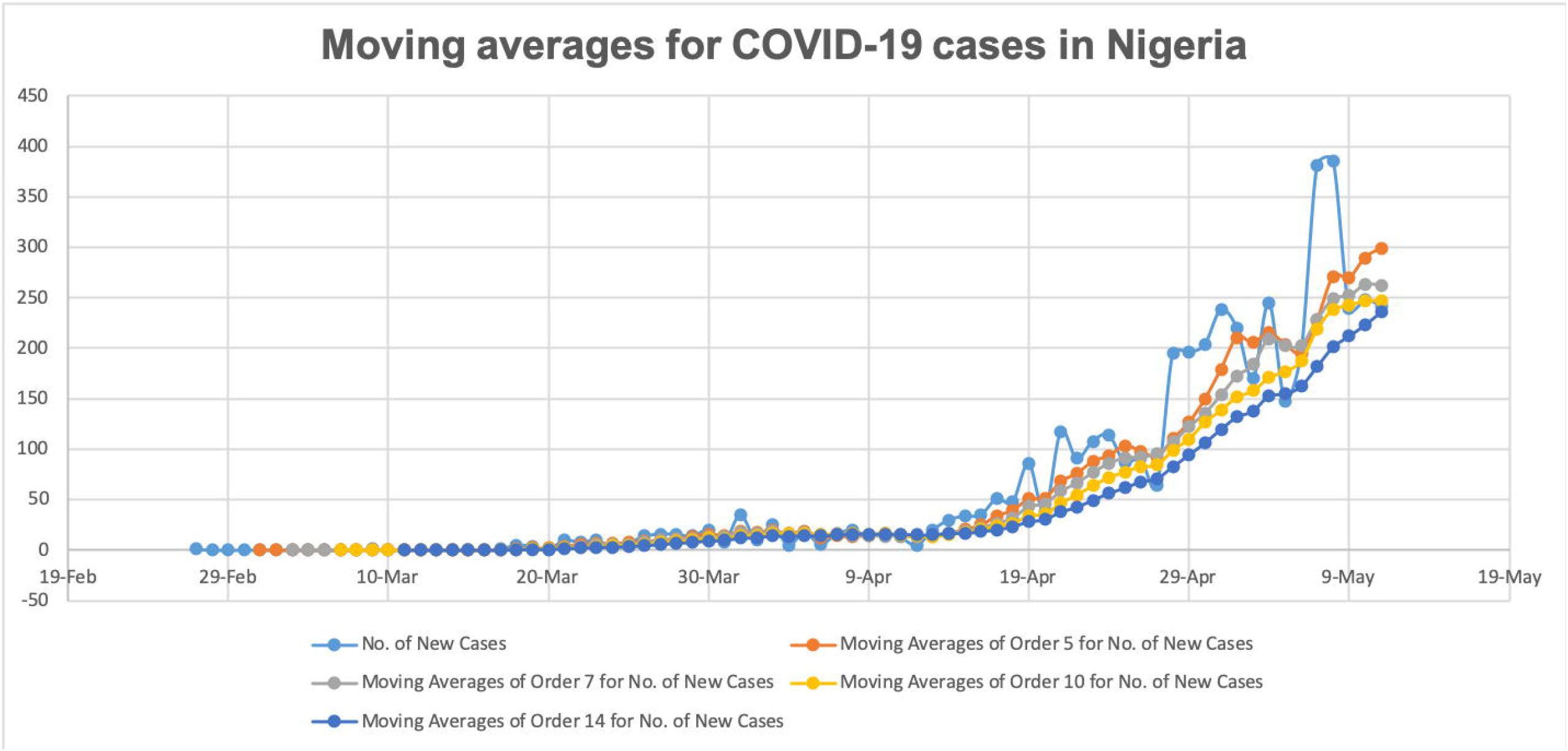
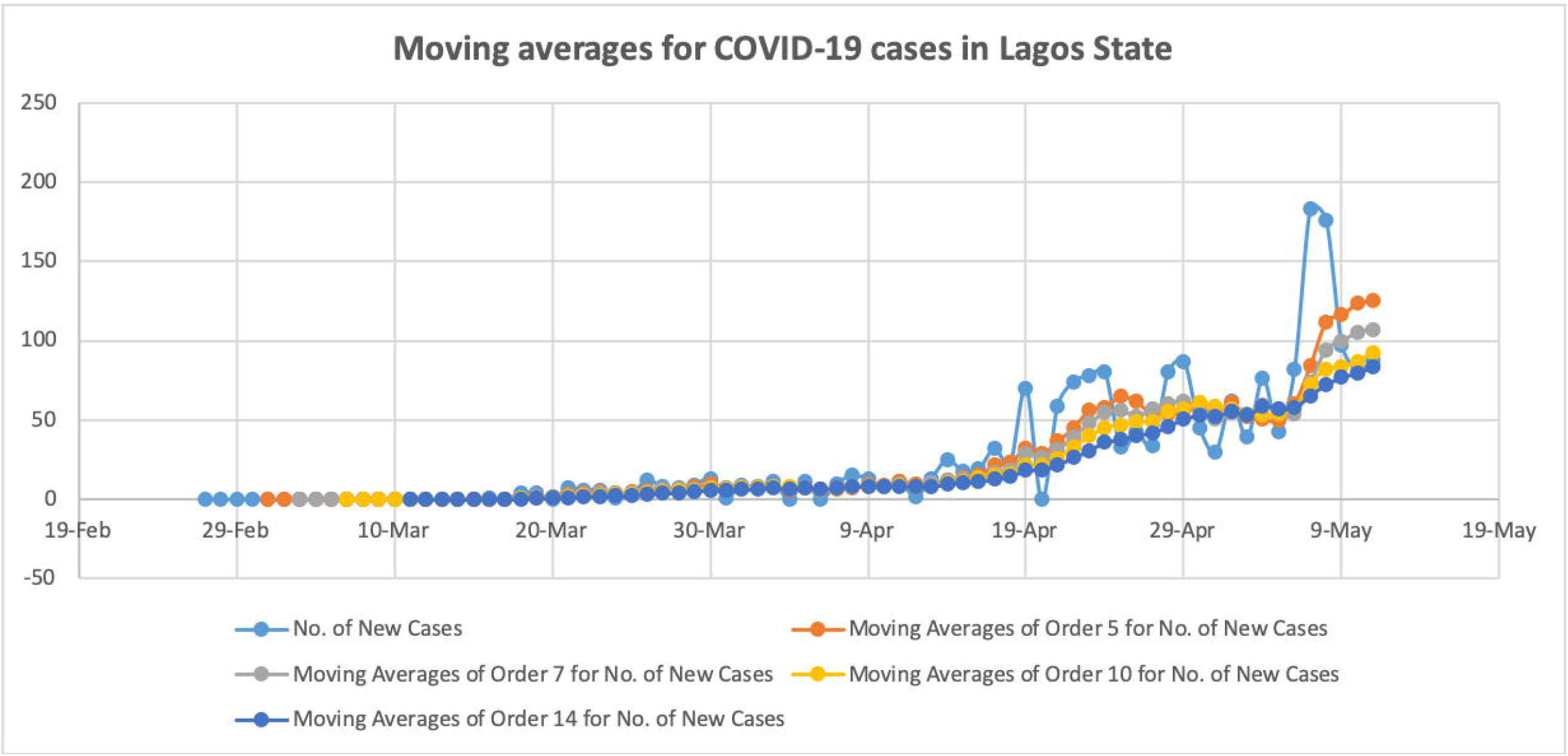
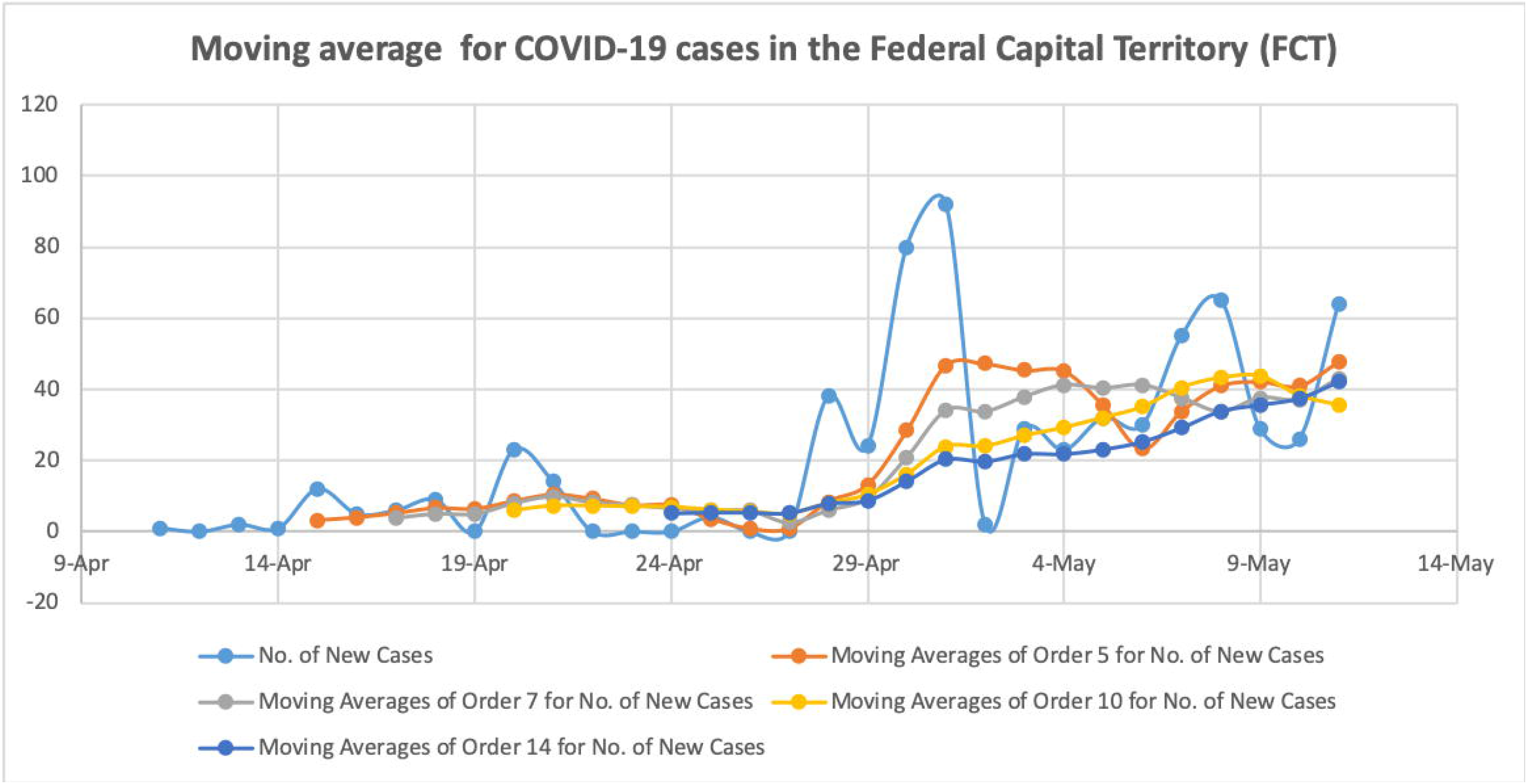
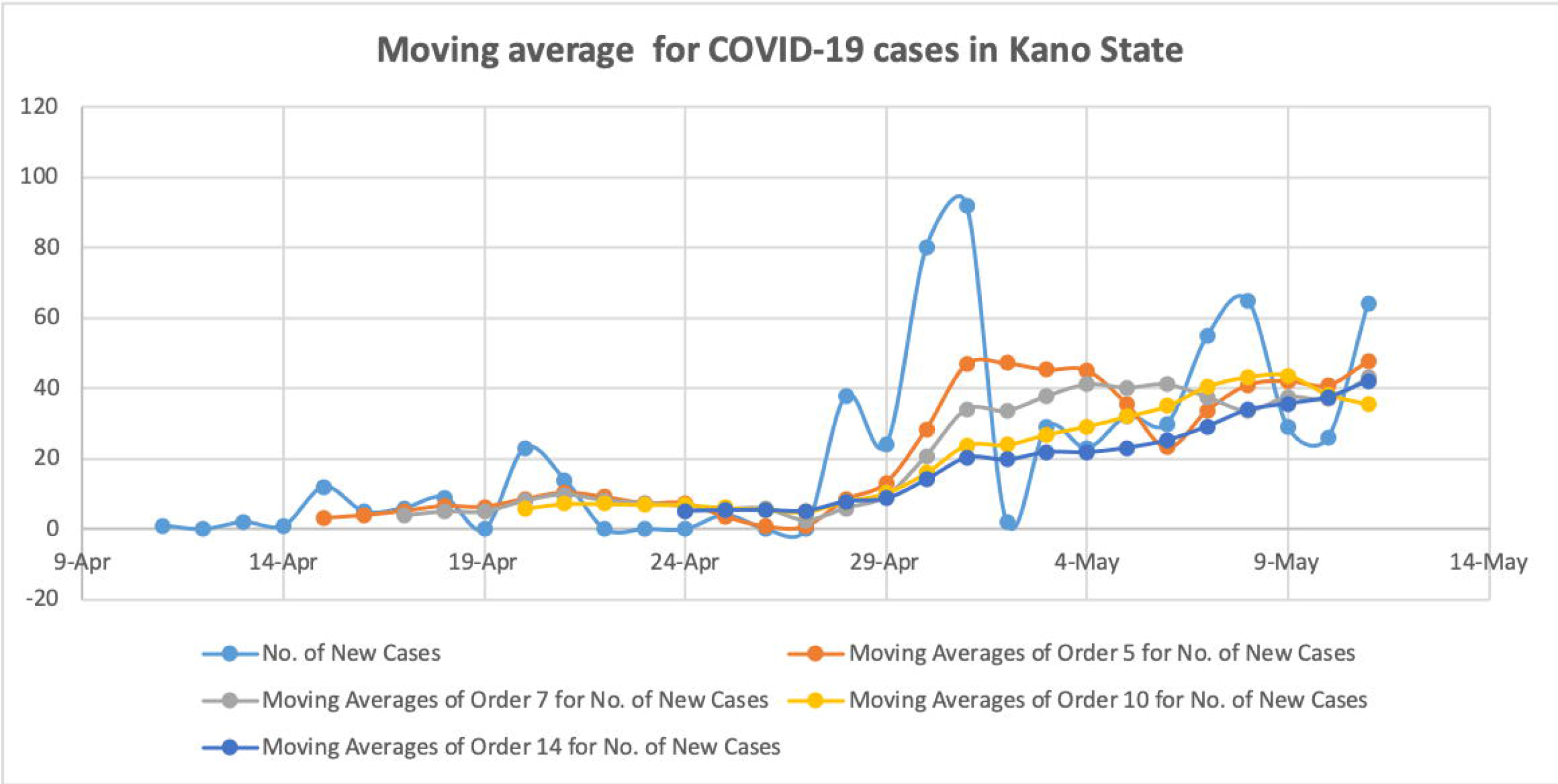
Moving averages of the COVID-19 cases in Nigeria, Lagos, Kano and the FCT

**Figures 5-8:**
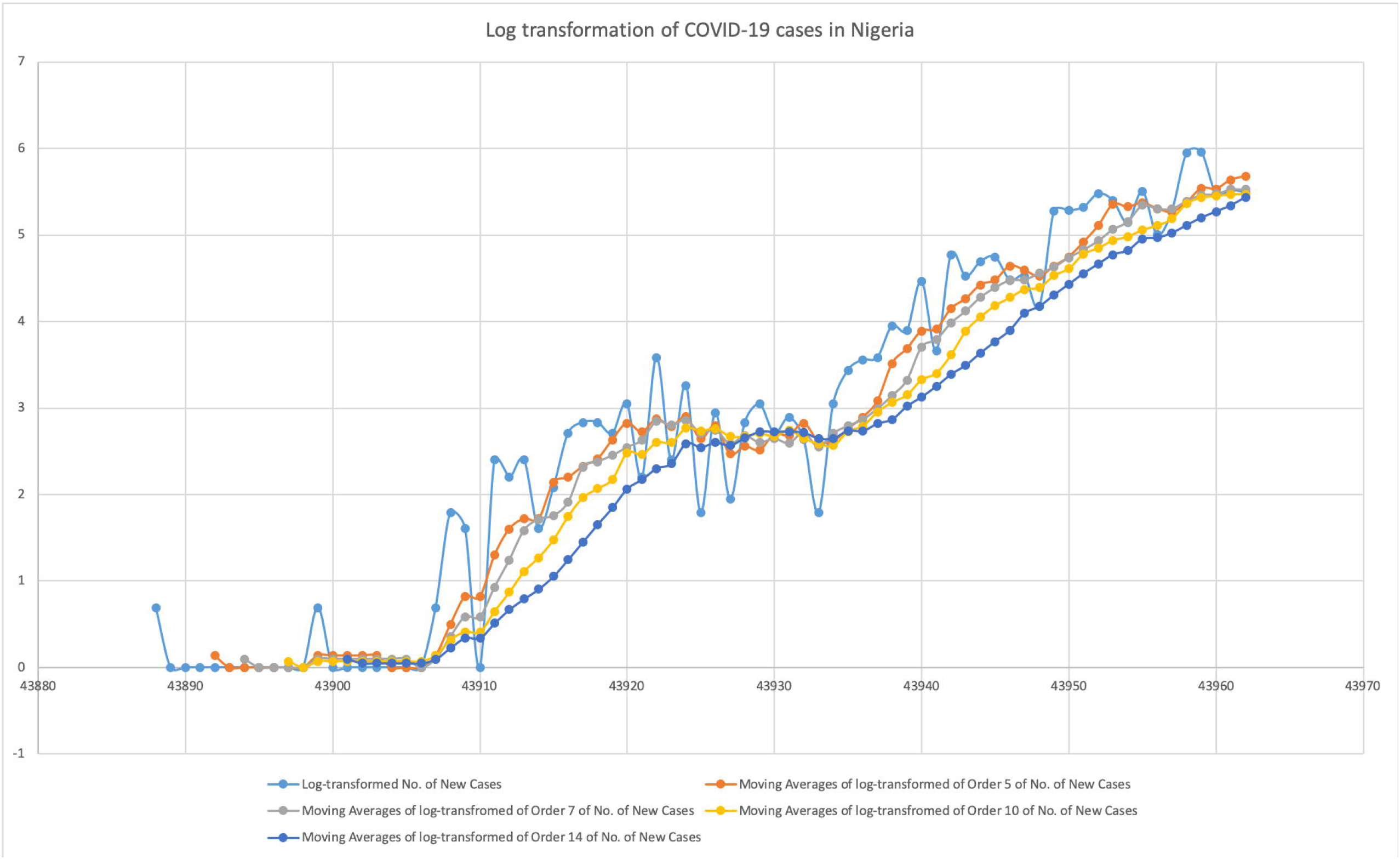
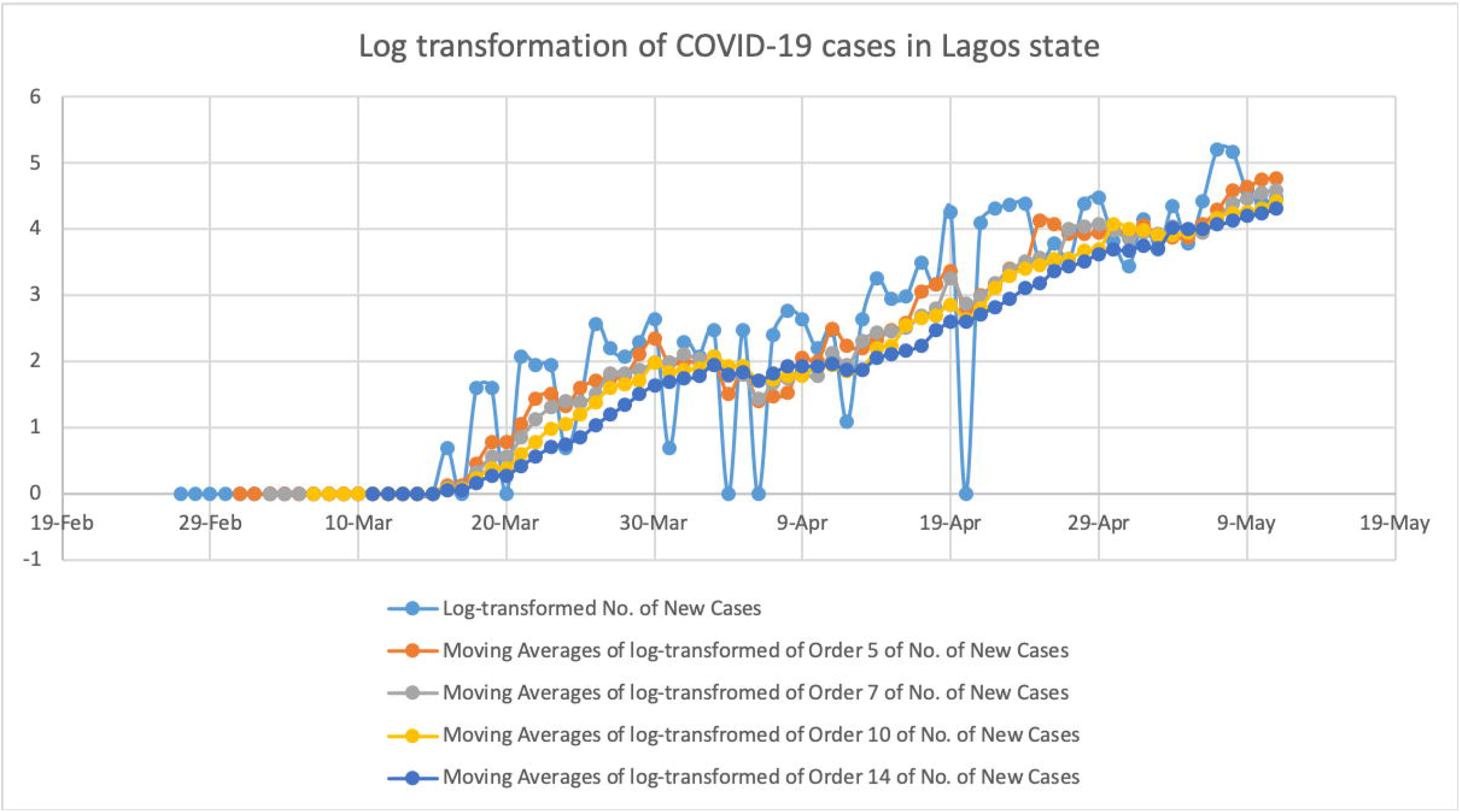
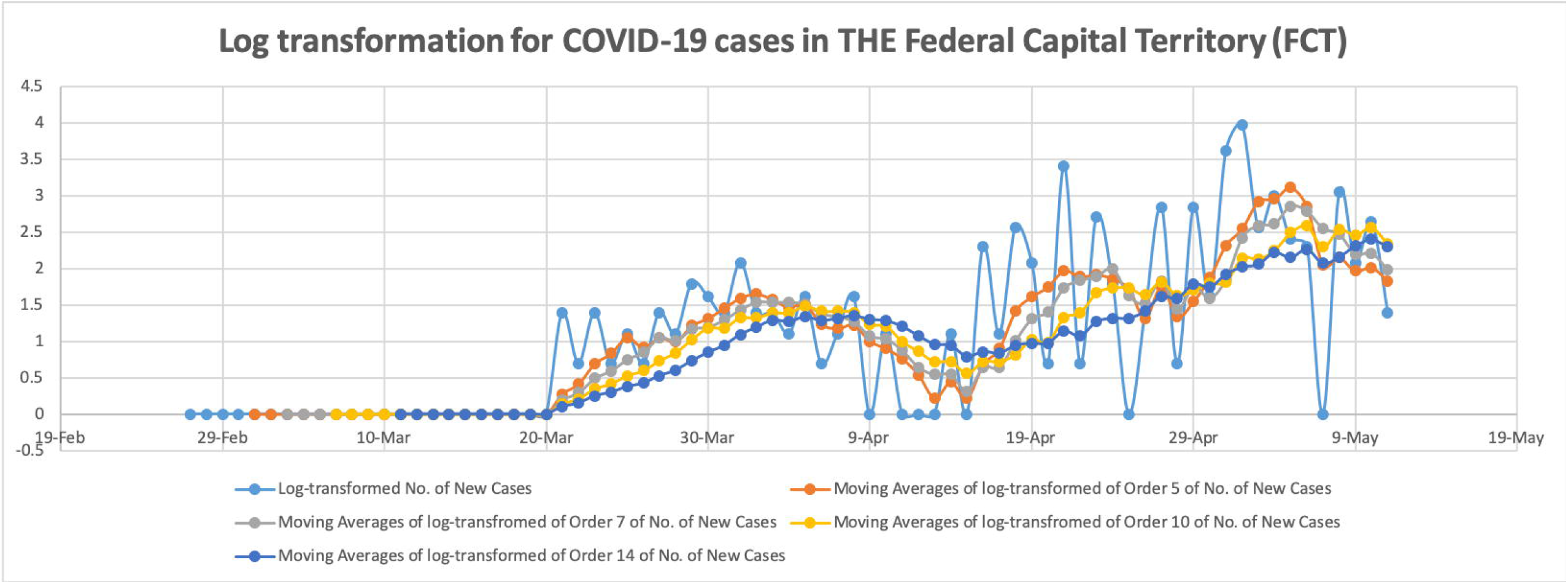
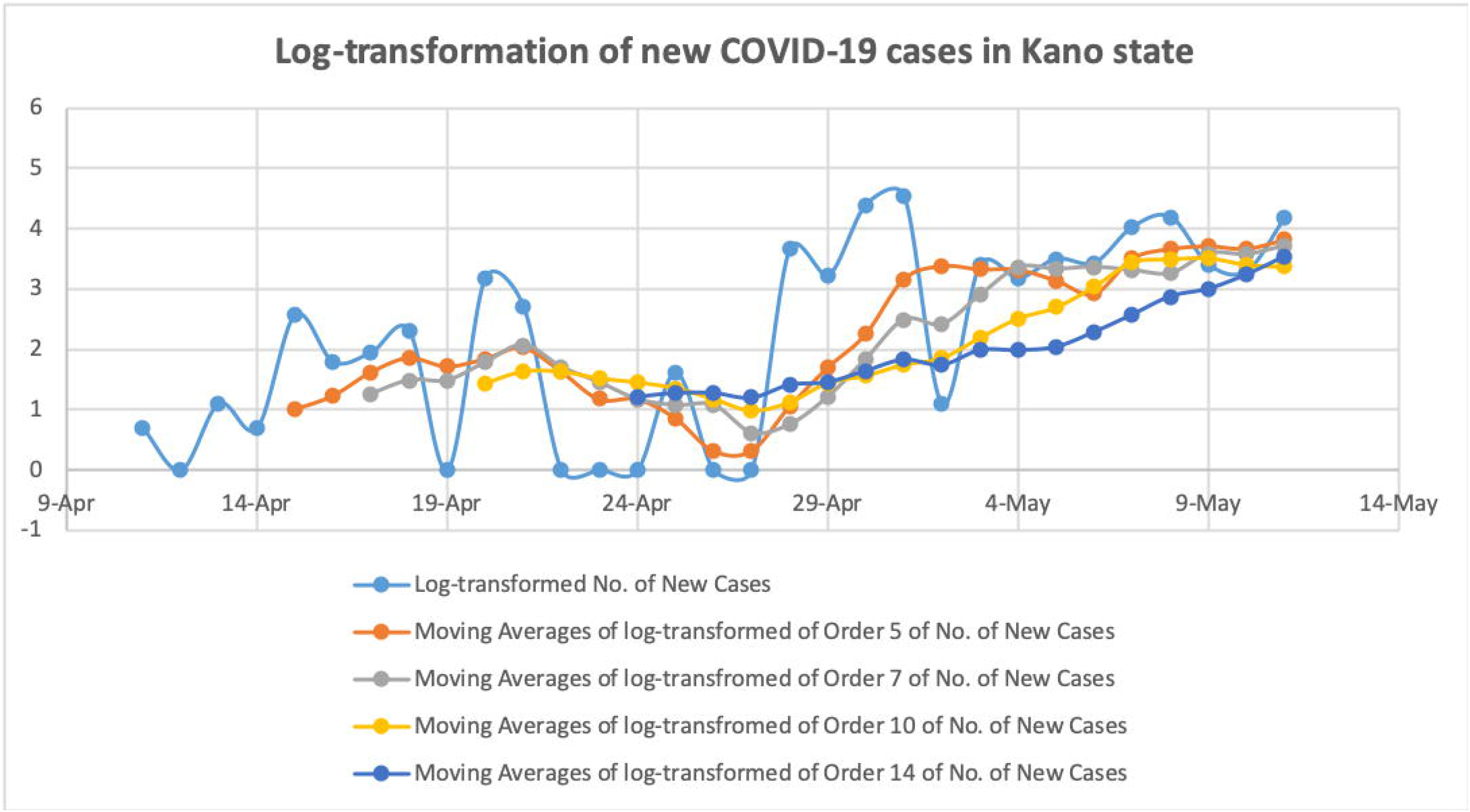
The log transformation of the moving averages of new cases of COVID-19 nationally and in Lagos, Kano and the FCT.

Figures 9-12 show the Epidemic Evaluation Index of the COVID-19 cases nationally and in Lagos, Kano and the Federal Capital Territory. The blue lines show the EEIs and the grey and red lines show the best and worst cases scenarios respectively. Nationally and in Lagos, the EEI started off on very high notes, however, the effects of the lockdown seem to be observed by the end of April and early May, as the EEIs are closer to 1. In all case scenarios, the EEIs are still above 1 indicating that despite the progress that has been made, the epidemic is still not fully under control and Nigeria has not yet reached its peak or entered a period of decline.

**Figures 9-12:**
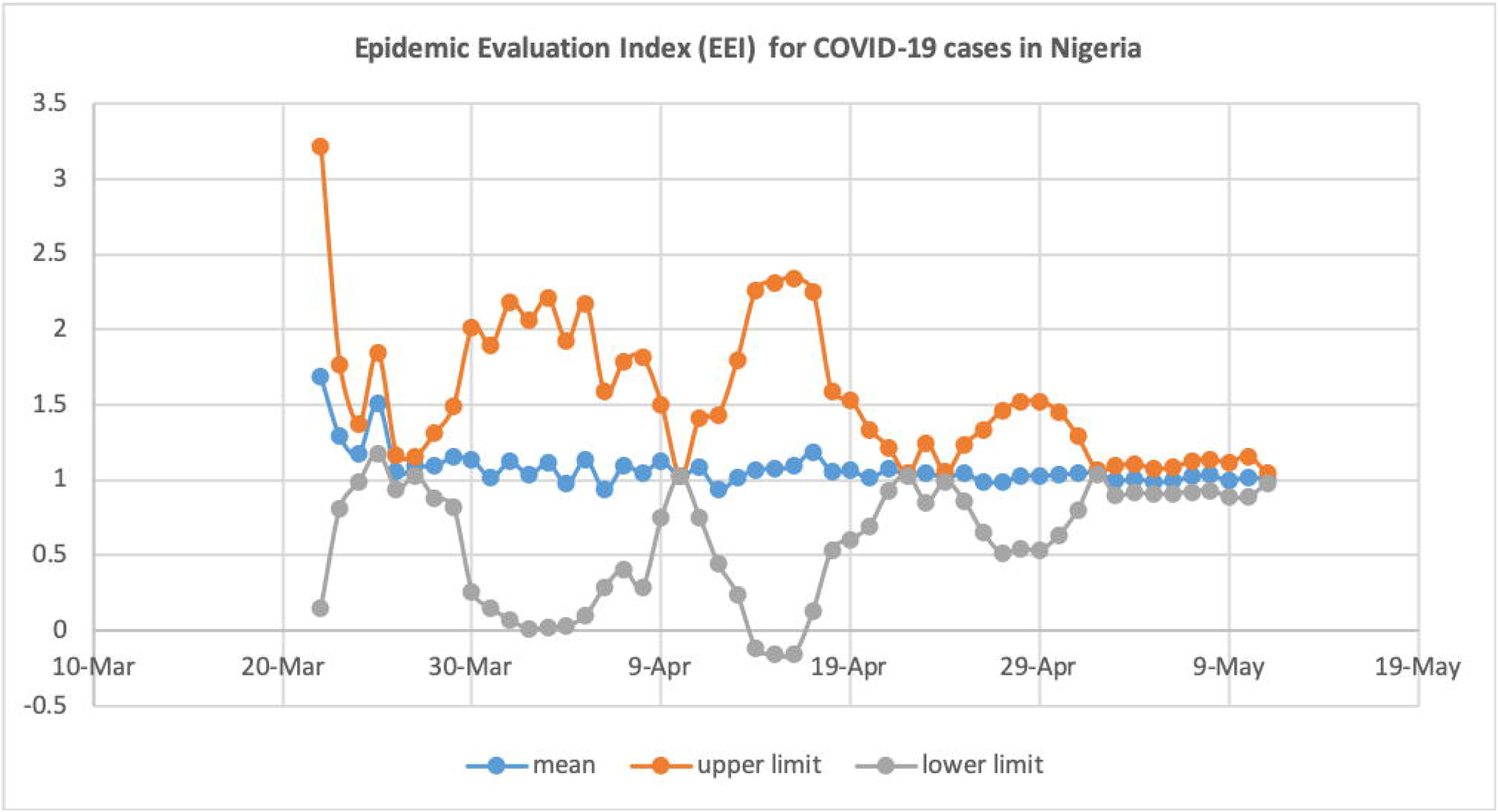
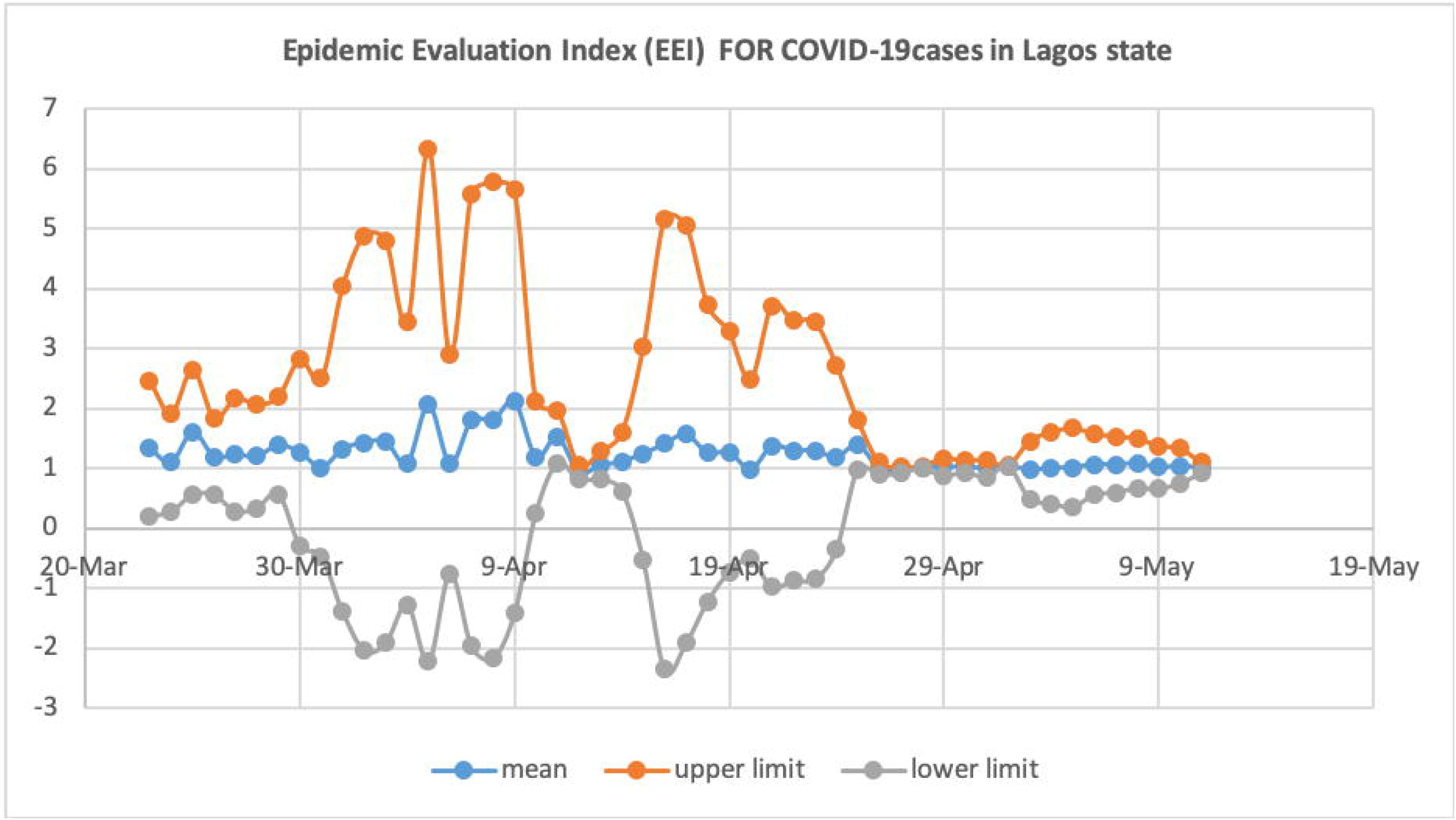
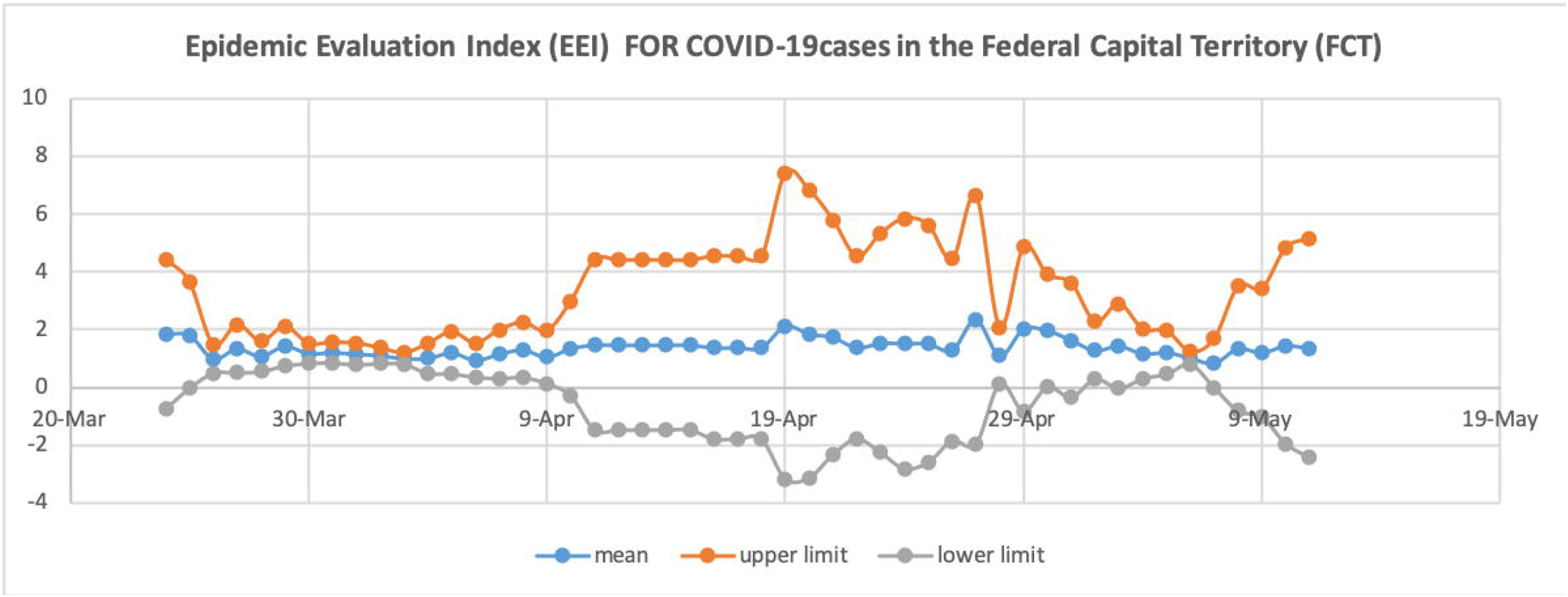
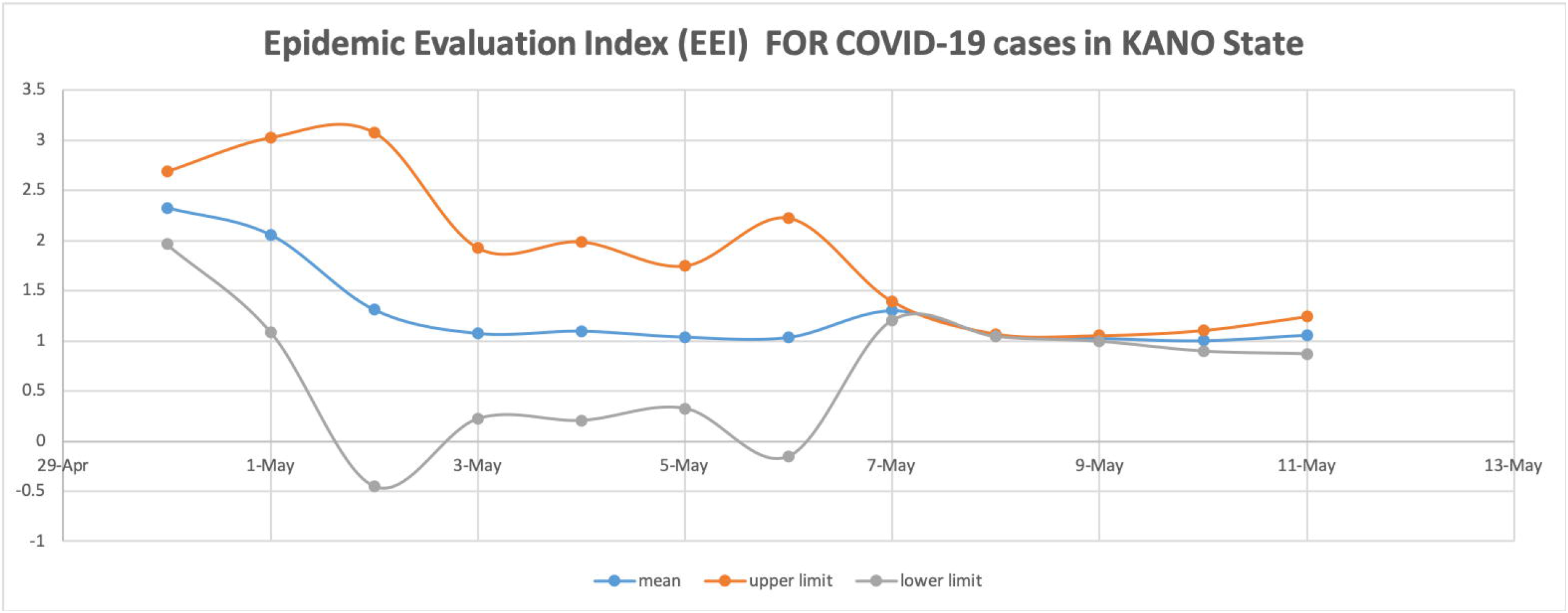
Epidemic Evaluation Index of the COVID-19 cases nationally and in Lagos, Kano and the Federal Capital Territory.

Tables 1 & 2 show that the EEIs are above 1 nationally and in the specific states. Table 2 shows that the EEI appears to have reduced after the lockdown with slight variations in the rates of reduction across states.

**Table 1.** shows the computed epidemic evaluation indices in Nigeria and specifically in Lagos, Kano and the FCT

| Epidemic Evaluation Index (EEI) |  |  |  |  |
| --- | --- | --- | --- | --- |
|  | National | Lagos | FCT | Kano |
| Mean | 1.0770 | 1.2344 | 1.3849 | 1.2811 |
| Standard Error | 0.0175 | 0.0388 | 0.0453 | 0.1272 |
| Median | 1.0454 | 1.1834 | 1.3562 | 1.0655 |
| Standard Deviation | 0.1252 | 0.2744 | 0.3172 | 0.4406 |
| Sample Variance | 0.0157 | 0.0753 | 0.1006 | 0.1941 |
| Kurtosis | 12.9649 | 2.2396 | 0.9136 | 2.5661 |
| Skewness | 3.2631 | 1.5140 | 0.9449 | 1.9129 |
| Minimum | 0.9366 | 0.9308 | 0.8431 | 1.0014 |
| Maximum | 1.6852 | 2.1076 | 2.3338 | 2.3261 |

**Table 2.** shows the Epidemic Evaluation Index before, during and after the lockdown in Lagos and the FCT.

| Epidemic Evaluation Index before, during and after the lockdown |  |  |  |  |
| --- | --- | --- | --- | --- |
|  |  | Before Lockdown | During Lockdown | After Lockdown |
| **National** | Mean | 1.24 | 1.04 | 1.01 |
|  | Min | 1.03 | 0.94 | 0.99 |
|  | Max | 1.69 | 1.19 | 1.03 |
| **Lagos** | Mean | 1.28 | 1.26 | 1.04 |
|  | Min | 1.09 | 0.98 | 1.01 |
|  | Max | 1.60 | 2.11 | 1.08 |
| **FCT** | Mean | 1.38 | 1.42 | 1.20 |
|  | Min | 1.00 | 0.95 | 0.84 |
|  | Max | 1.84 | 2.33 | 1.44 |

## Discussion

Globally, effective strategies are required to prevent and control the spread of the COVID-19 pandemic. Estimating the evolutionary trends of this outbreak is crucial for the allocation of scarce medical resources, regulation of human activities, and even for the national economic development of countries. Thus, the creation of a reliable and suitable model to help governments decide on emergency macroeconomic strategies and medical resource allocation is crucial.^6^

This paper describes the epidemiological trends of COVID-19 cases in Nigeria as a whole and in the two most affected states i.e. Lagos and Kano as well as the FCT. At the national level, our analysis show that the COVID-19 epidemic curve is rising steadily but yet to reach its peak. We observed that the direction of the moving averages of different states in Nigeria are rising. It is evident that the rate of growth of the logarithm-transformed moving average had slowed during the lockdown.

A computed EEI of greater than 1.0 suggests that the epidemic is still evolving and this means that aggressive public health restrictions must be sustained. If the EEI drops below 1.0, it can be judged that the epidemic has entered the period of decline, meaning that it may be time to plan to phase out some of the stringent preventive and control measures. If the EEI remains below 1.0 for more than 7 days, it indicates that the epidemic has entered a stable period of transmission. In Nigeria, the all the computed EEIs are yet to fall below 1.0 indicating that the epidemic in Nigeria is still evolving, highlighting the need to maintain aggressive public health measures.

Since the first recorded case of COVID-19 in Nigeria, several public health measures have been instituted in a bid to control the spread and limit the number of new infections. Between March 18 and 30 2020, the Federal Government closed all educational institutions, placed restrictions on large social gatherings and air travel and land borders and began an aggressive awareness campaign to promote public health behavioural change. Our study shows that some of these stringent measures have had some positive effects on the transmission rates. However, we observe that active transmission is still on-going and these measures may need to remain in order to further reduce transmission rates.

Mathematical models can help to identify the best timing for government intervention measures as seen in China, where peak of the epidemic was predicted with a model showing that aggressive control measures caused a decline in the number of new cases.^9^ Similar models have effectively informed public health decision making. For instance, in Lima, Peru, investigators used a generalized growth model to forecast an early exponential growth trend in Lima, which slowed down and turned into an almost linear growth trend, highlighting the need to continue social distancing and active case finding efforts in the region.^10^ In India, authors showed a short-term trend of COVID-19 for the three highly affected states, Maharashtra, Delhi, and Tamil Nadu, and suggested that the first two states needed further monitoring of control measures.^11^ In Italy, France and Spain, the auto-regressive integrated moving averages (ARIMA) models were developed and showed an initial exponential trend of new cases with a gradual decline as government planned to return to normal life. Meanwhile, Spain, Europe’s second-worst-hit country at the time saw a trend that suggested a drop in daily coronavirus deaths though the number of confirmed cases had overtaken Italy. In France on the other hand, similar to our observations, no downward trend was observed at the time of the study, indicating that France, like Nigeria, had not yet reached its epidemic peak.^6^

One of the principal steps taken by several governmental agencies to control COVID-19 is the enactment of lockdowns to maintain social distance. This procedure is an excellent measure to control the spread of the disease. Still, from an economic point of view, complete lockdowns may be the cause of a significant financial crisis for the near future.^11^ Consequently, competing economic priorities may make a full lockdown for an indefinite period undesirable.^11^ However, lockdowns in densely populated states like Lagos and Kano, may reduce the disease transmission rate, although complete control may not be achievable. Therefore, a suitable balance of governmental policies is often preferable. Our data-driven findings are relevant for decision makers as they attempt to strike this balance.

To the best of our knowledge, this study is one of the first to employ moving averages and EEI models to evaluate the trends of COVID-19 in an African setting and in Nigeria in particular. It however has some limitations, Firstly, moving averages are often affected by extreme values and outliers, however, our data did not have significant outliers or extreme values. Secondly, logarithmic transformations of the moving averages might be problematic when several zero entries on consecutive days producing an average of zero exist in the dataset. This was taken care of by adding 1 to the moving averages before computing the logarithms. Another limitation is that our EEIs were computed based on a singular moving average order 7. There may be a need to consider the comparison of other orders to check for consistency in future studies. Further, the data used to develop our models are based on publicly available data from the NCDC. As at the time we constructed our charts, only 29,408 tests had been conducted nationally, indicating a possible under estimation of our findings. Also, the limited data from Kano (the first case was on the 11^th^ April 2020) may make inferences for Kano a little premature. Finally, all data presented are as at the 12^th^ May 2020, events after this are not included in our model, however, we hope to report an update of our findings over time.

### Conclusion and recommendations

The number of new cases of COVID-19 has been on a steady rise since the first reported cases on the 27^th^ February 2020. Public health interventions including the lockdown measures in Lagos and the FCT appear to have assisted in reducing the rate of increase of new infections. In Nigeria and across the two states and the FCT, the EEIs are still above 1, suggesting that the epidemic in Nigeria is still developing and has not yet reached its peak. We recommend that aggressive public health interventions and restrictions against mass gatherings should be sustained.

## Data Availability

The data used in this article is from public sources and available at www.ncdc.gov.ng

## Acknowledgements

The protected time for the contribution of OO (Oluwakemi Odukoya) towards the research reported in this publication was supported by the Fogarty International Center of the National Institutes of Health under the Award Number K43TW010704. The content is solely the responsibility of the authors and does not necessarily represent the official views of the National Institutes of Health.

